# Patients experience with preoperative use of anti-obesity medications and associations with bariatric surgery expectations

**DOI:** 10.1101/2024.04.14.24305798

**Authors:** Jason M Samuels, Mayur Patel, Christianne L. Roumie, Wesley Self, Luke Funk, Matthew Spann, Kevin Niswender

## Abstract

**Objective:** This study investigated associations between patients’ experiences with anti-obesity medications and weight loss expectations prior to bariatric surgery.

**Methods:** Patients were electronically surveyed with a 31-item questionnaire via email or the patient portal with a primary predictor variable of AOMs pre-surgery. Outcomes included degree of weight loss and weight regain and motivation for seeking surgery.

**Results:** 346 persons were invited to complete the survey. 112 (32.4%) were completed, with 7 excluded due to not answering the AOM question. 73% reported AOM use. Among those who took AOMs prior to seeking bariatric surgery, average weight loss was 13 kg (SD ±10 kg) corresponding to a 4.4 kg/m^2^ decrease in BMI. Of past AOMs receipients, 87% reported weight regain upon stopping AOMs. Average weight regain was 18 kg (SD 13kg, 126% increase). Patients reported improved longevity and quality of life as motivation for seeking surgery with AOM use history having no effect. Subjects reported an average weight loss goal of 65.8 kg (39% of baseline weight) from bariatric surgery.

**Conclusion:** AOMs were commonly used in those seeking bariatric surgery but motivation for surgery did not differ by AOMs use history. Motivations were most often related to goals for better overall health.

**Study Importance Questions:**

- Minimal data exists regarding patients’ experience with anti-obesity medicaitons and whether prior experience with these medications impacts patients’ expectations or goals for bariatric surgery.
- This cross-sectional survey investigated the frequency of anti-obesity medication use prior to patients seeking bariatric surgery, finding nearly 3/4ths of patients seeking surgery have previously utilized pharmaceutical weight loss therapies.
- This study will ideally spur clinical trials to determine the most effective approach to combination therapy, thus moving obesity treatment to a multimodal approach.

## Introduction

As the therapies targeting obesity expand, patients’ experiences with weight loss treatments continue to evolve. Several medications for weight loss were approved by the Food and Drug Administration (FDA) in the last 5 years, including the latest generation of Glucagon-like Peptide 1 receptor agonists (GLP1RA) which achieve 15-20% total weight loss (TWL).(1, 2) Most recently, in 2023 the FDA approved Tirzepatide, a glucose-dependent insulinotropic polypeptide (GIP)/GLP1RA di-agonist, based on the results of the SURMOUNT 1 trial in which participants lost on average 21% of their body weight. Building on the interest in prior GLP1RA therapies approved for weight loss such as semaglutide, the resulting FDA approval of Tirzepatide for weight loss received considerable media coverage.(3, 4) As a result, these latest generation of AOMs with GLP1RAs and GIP/GLP1RAs represent the fastest growth in prescribing of all pharmaceuticals and represent a landmark change in the approach to obesity treatment.(5, 6)

Despite these pharmaceutical advances, bariatric surgery remains the most effective treatment with TWL averaging 25-35% of pre-surgical body weight.(7, 8) Prior studies demonstrate that patients’ expectations for weight loss after surgery exceed the average achieved with surgical weight loss alone.(9) Cohn reported that patients seeking surgical weight loss are motivated by a desire to improve overall health or quality of life, rather than to achieve a particular weight.(10) These studies largely predate the use of medications specifically developed and approved for treating obesity. Thus, little is known about the frequency of use of AOMs prior to bariatric surgery, or how the growth of AOM use has impacted patients’ motivations and expectations for bariatric surgery.

Our aim was to describe the frequency of AOMs prior to seeking surgical weight loss. We also sought to evaluate the association between patients’ experience with AOMs and weight loss expectations prior to bariatric surgery, the motivations for seeking surgical weight loss and desired weight loss.

## Methods

### Study design and Survey Instrument

This single-center, cross-sectional survey study was conducted at the Vanderbilt University Medical Center Surgical Weight Loss Clinic between December 13, 2023, and March 13, 2024. The survey instrument consisted of 31 items (Appendix 1). Domains included: Demographics, AOMs used, Motivation for seeking weight loss treatment, and Items regarding combination weight loss strategies. Patients were asked to rank their selected motivatons on a scale of 1 to 6 with 1 being least important and 6 being most important. Remaining items were multiple choice, Likert-scale, and free text responses. Completion of the survey required 5-10 minutes, and participants did not receive compensation for completion. The institutional review board deemed this study exempt from IRB review (IRB #232013), and consent was implied with completion of the questionnaire.

### Study Population

We identified eligible participants from the clinic schedule if they were listed as a “new consult” patient who was seeking bariatric surgery, age ≥18 years old, English speaking, and had an active My Health at Vanderbilt (MHAV) patient portal account or email on file. Exclusion criteria included those patients listed as “do not contact” for research studies and those who did not answer the item regarding prior AOM use. Participants who noted prior bariatric surgery were eligible for inclusion.

### Instrument Administration and Data Collection

The instrument was administered electronically via the patient portal application, a patient-centered mobile app with messaging capabilities, or via email. The message contained information about the survey and a link to complete the survey electronically. All participants received a reminder message to complete the survey within one week of the initial message. No participant data was collected beyond the information participants provided within the survey instrument.

### Predictor Variable: AOM

The primary predictor was patient reported usage of AOMs at any point prior to their visit to the surgical weight loss clinic. Any use, as determined by the patient, was considered as an exposure to AOMs. A complete list of medications, including generic and brand names, can be found in Appendix Table 1. Among those who used AOM, we describe their self-reported weight loss and weight regain.

### Outcomes: Motivation for Weight loss and desired weight loss goal

We compared the association between patients’ motivations for undergoing bariatric surgery and goals among those with and without use of prior AOM. Possible motivations used in the survey instrument were derived using commonly reported motivations from previously published literature.(11–14) The secondary outcomes included patients’ self-reported desired weight loss from surgery.

### Statistical Analysis

Data was collected using REDCap.(15) Descriptive statistics were utilized to report survey responses. Categorical variables were presented as frequency and percentage, and continuous variables were presented as median and interquartile range or means and standard deviation as appropriate. Medians (with interquartile range) and percentages were compared using the Wilcoxon-Mann-Whitney test, chi-square test, or Fisher’s exact test as appropriate. A two-sided p value of <0.05 was considered statistically significant. Statistical analyses were conducted using GraphPad Prism Version 10.1.1, (Boston, MA).

## Results

### Study Cohort and Demographics

A total of 346 patients were invited to participate (279 by MHAV patient portal message and 67 by email), and 112 responded (32%). Response rates differed between those contacted by patient portal versus email (36% vs. 16%). Seven patients were excluded for non-response on the AOM item, providing a final cohort of 105 respondents. Median age of respondents was 48 years (IQR 42 - 59 years) with 77% of the cohort identifying as female. Respondents were predominantly White/Caucasian (63%) followed by Black/African American (20%), “other” (2%) and either American Indian or Native Hawaiian or other Pacific Islander (1.1% each). Eighteen patients (17%) reported having previously undergone bariatric surgery (**Table 1**). 73% reported having previously taken AOMs previously.

**Table 1:**
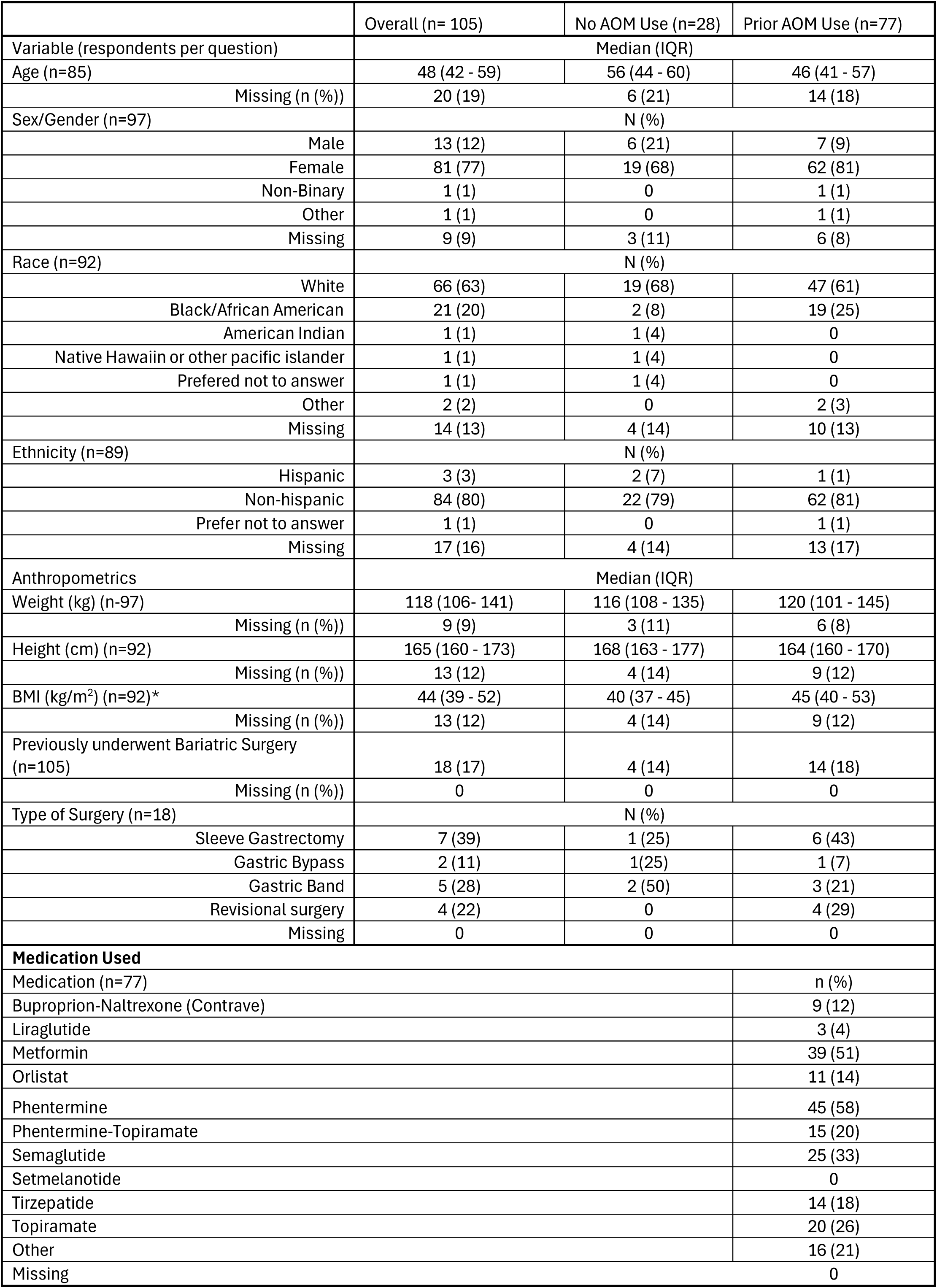
Baseline Demographics for overall cohort and the cohorts with histoy of AOM usage and without AOM usage.

Patients reported an average weight loss of 14.2 kg (± 10.9 kg) with prior AOMs use (median BMI loss with meds was 4.4 kg/m^2^). Sixty-five (87%) subjects reported weight regain after stopping the medication (two non-respondents), with average weight regain of 18.0 kg (± 12.9 kg, 126% of original weight loss).

### Participants Weight Loss Preference

Participants expressed a preference towards surgical intervention over medical intervention. In response to the item “If weight loss were the same, which approach to losing weight would you prefer to take?”: 43% selected surgery; 23% selected medications; and 33% were uncertain or had no preference. The majority of patients (73%) reported a “moderate” or “complete” willingness to take medications to enhance weight loss after surgery. Acceptance decreased to 58% if lifelong medication was require; or if the medication was a weekly injection (59% “moderate” or “completely” willing).

Participants were also asked “In which of the following scenarios would you be willing to take a weight loss medicine after bariatric surgery if the medicine is given as an injection taken weekly at your home?” Most respondents noted a relatively small amount of additional weight loss was necessary to consider such therapies. Participants expressed the greatest interest in adjuvant AOMs if they offered 20 lbs (35%) or 30 lbs (36%) additional weight loss. There were 30% who noted 40 lbs, 17% for 50lbs and 19% for 60 or more pounds of additional weight loss as needed to take an injectable AOM after surgery.

### Weight Loss Motivations and Expectations

Participants were asked to select their motivations for weight loss from a list of common reasons. Decreasing the risk of serious health problems was the most common answer (91%) followed by longer life expectancy (87%) and improved physical activity (82%). Achieving a lower weight was similarly, but not the most common (73%). When comparing participants with or without prior history of AOM usage, there was no difference between groups in the frequency of noted motivations for seeking surgical weight loss (all p > 0.1, Figure 1). We also compared how the two groups rated the importance of individual motivations. Only a desire to reduce pain differed in importance between groups, with patients who had no prior history of AOM usage rating pain reduction with greater importance than those with no AOM usage history (median 4 vs 2, p=0.003).

**Figure 1:**
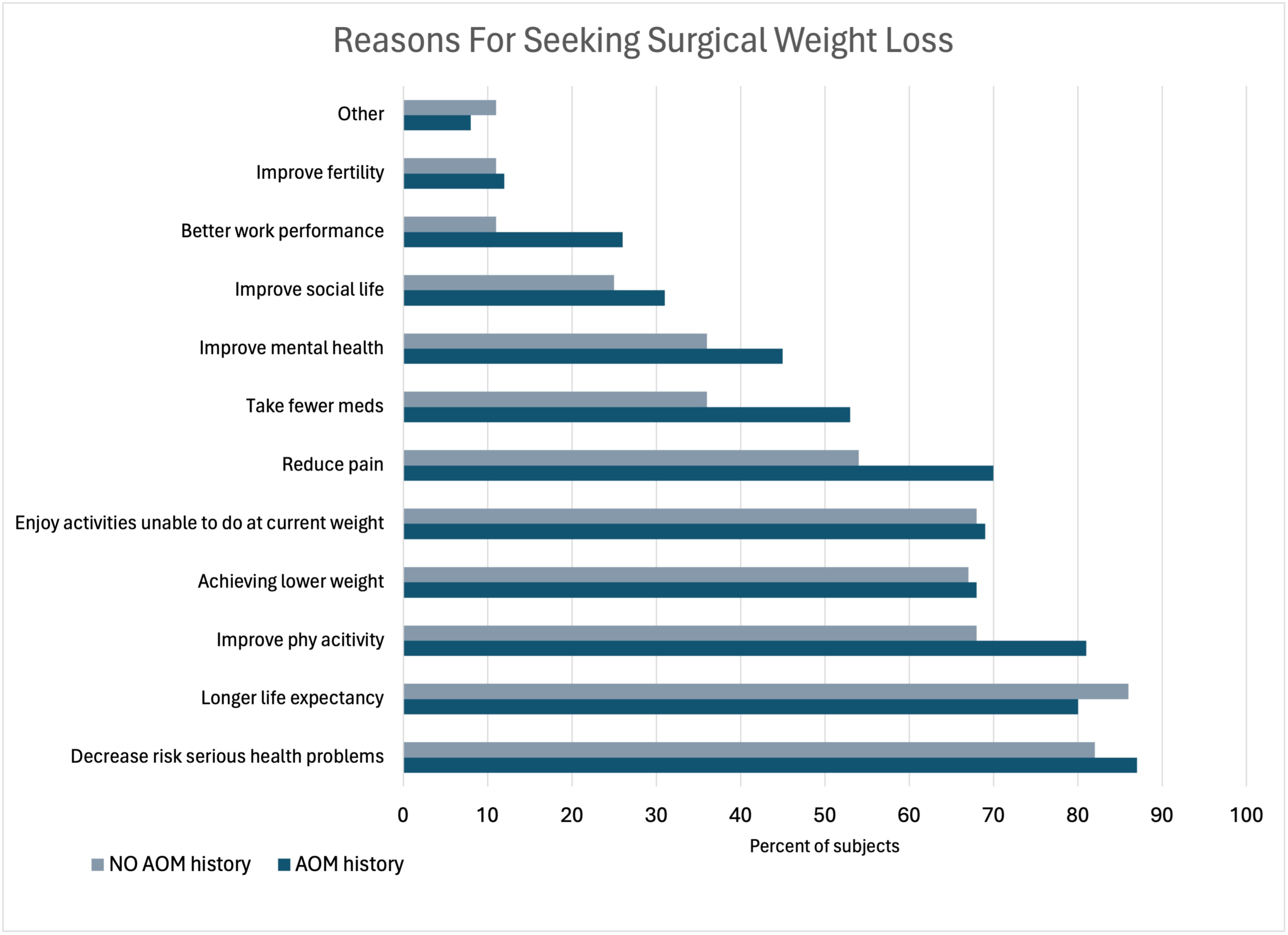
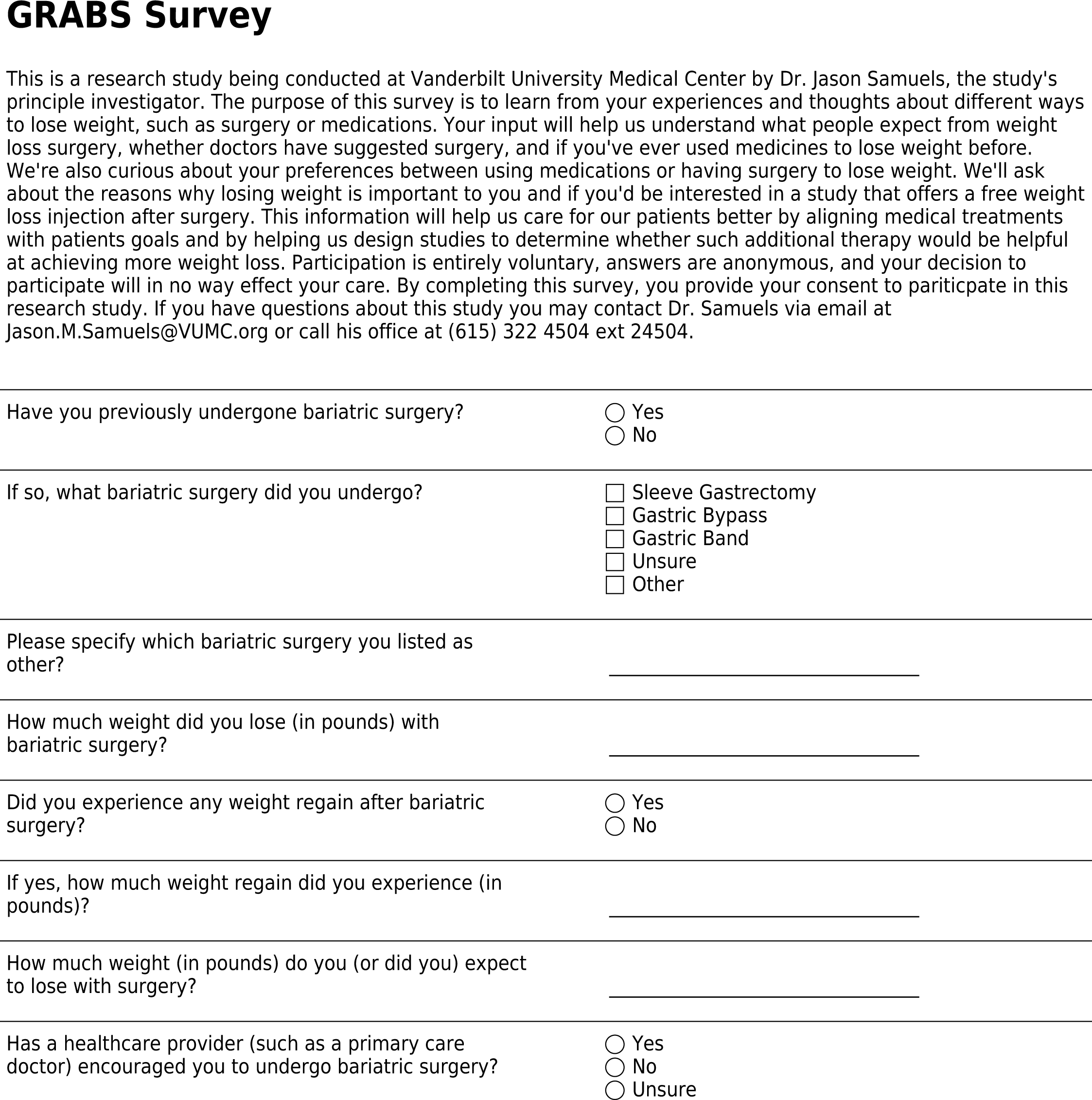
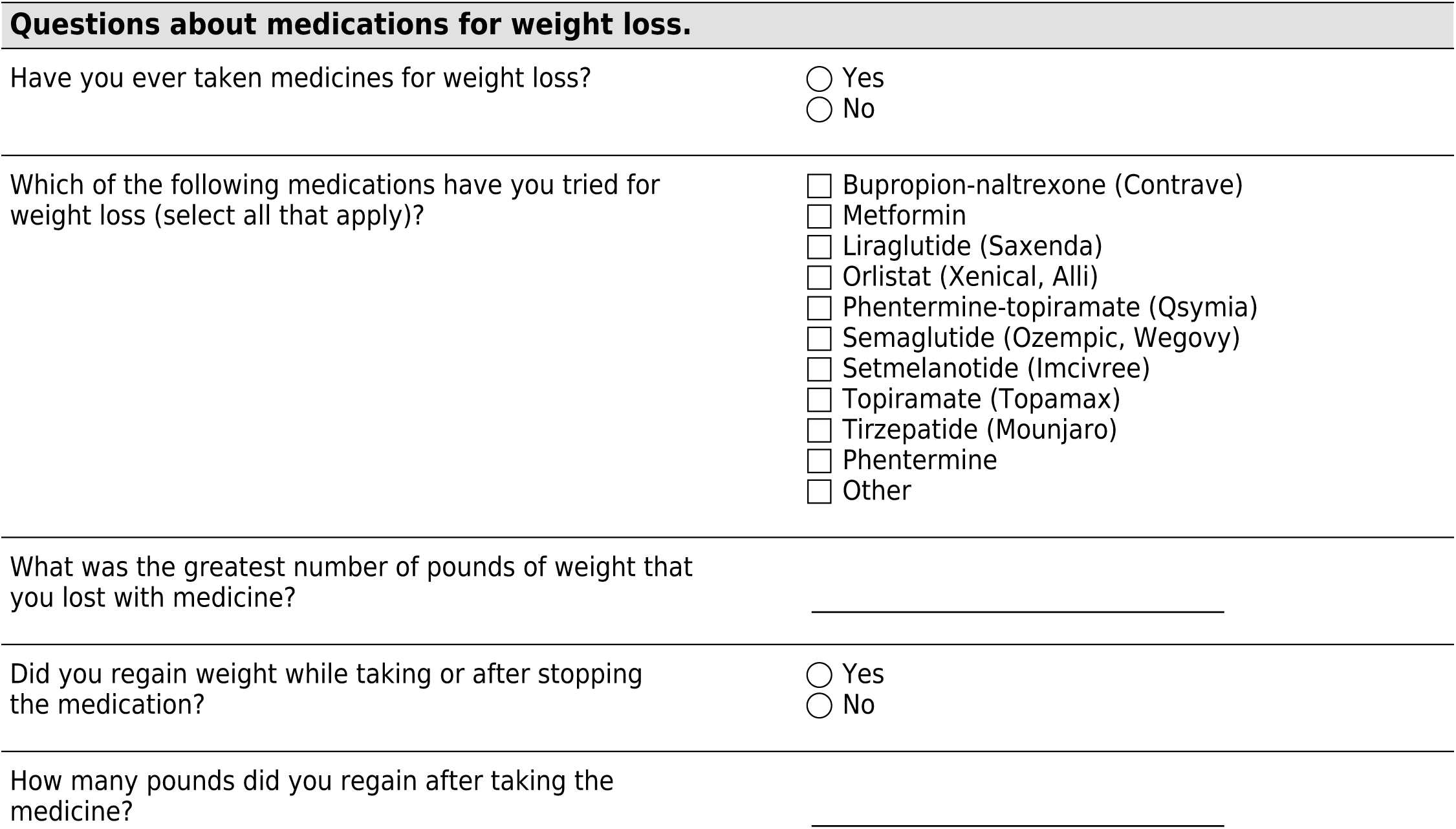
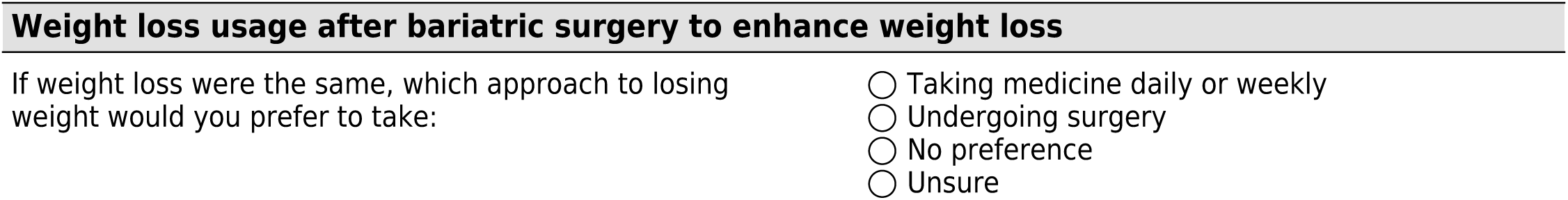
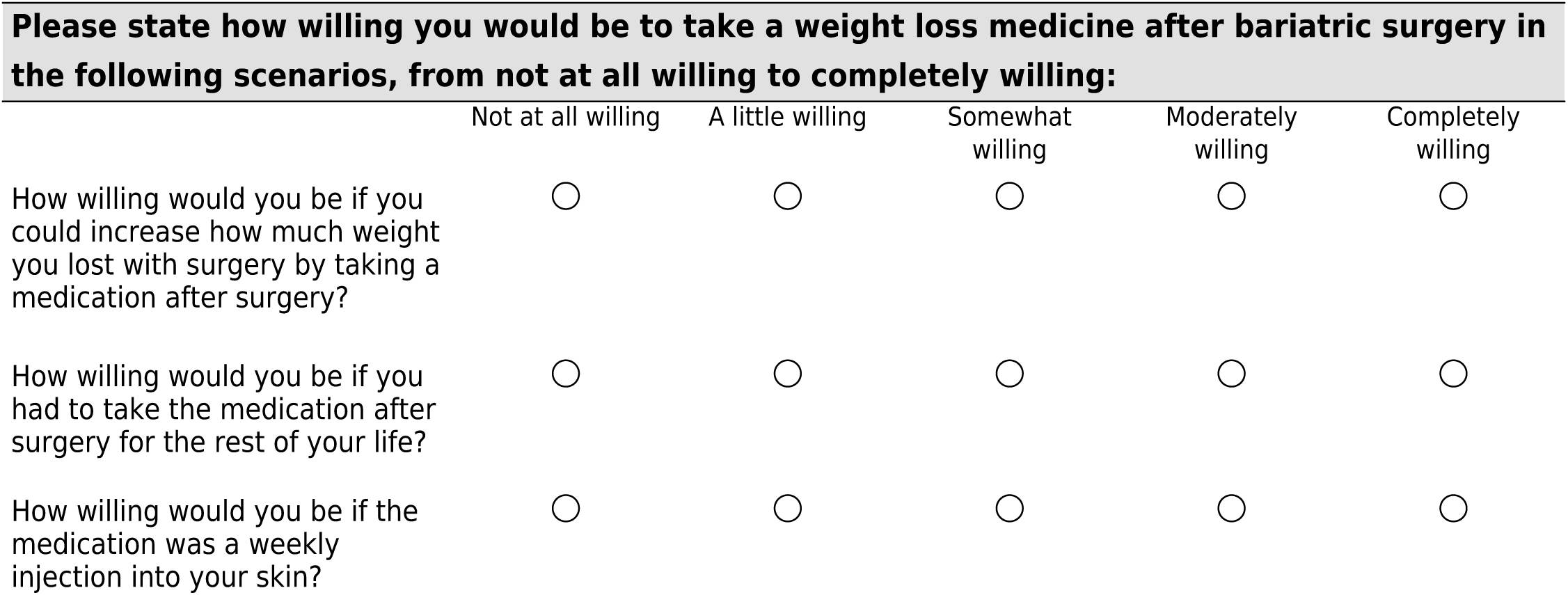
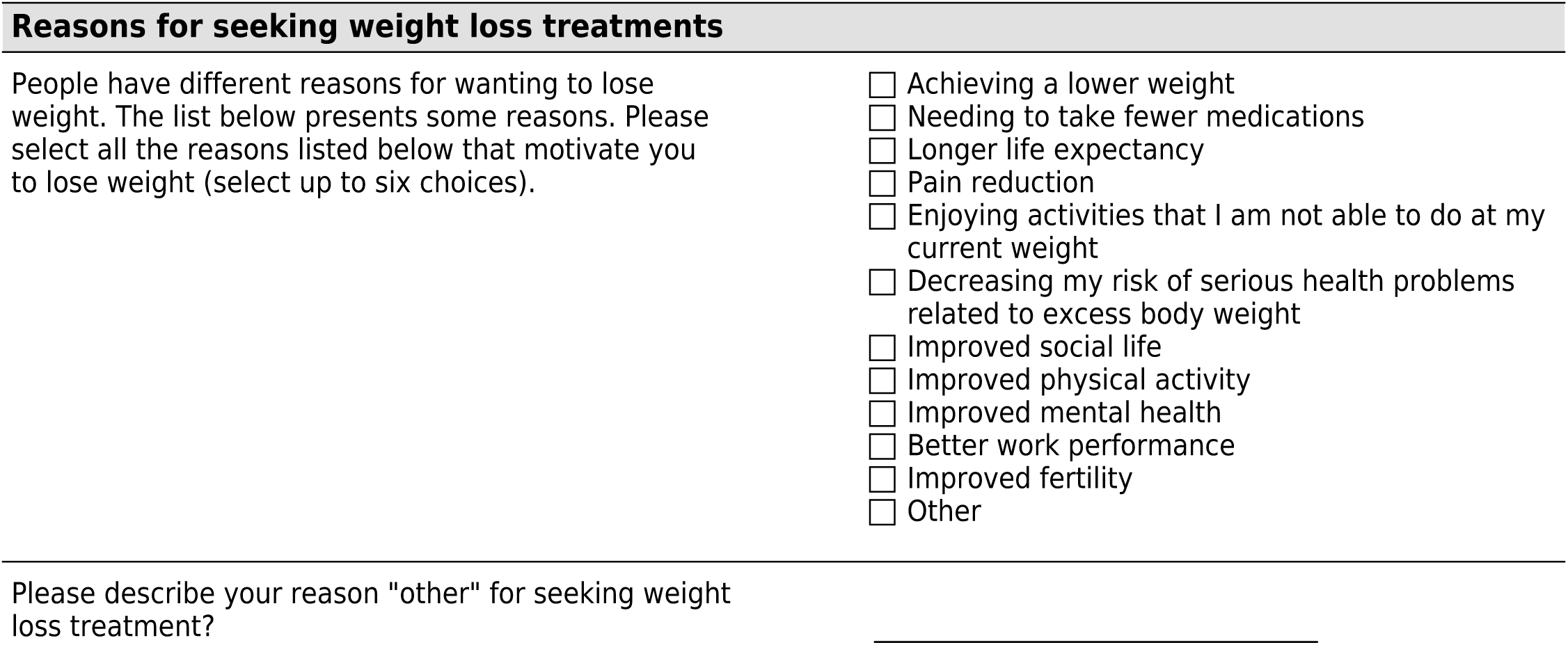
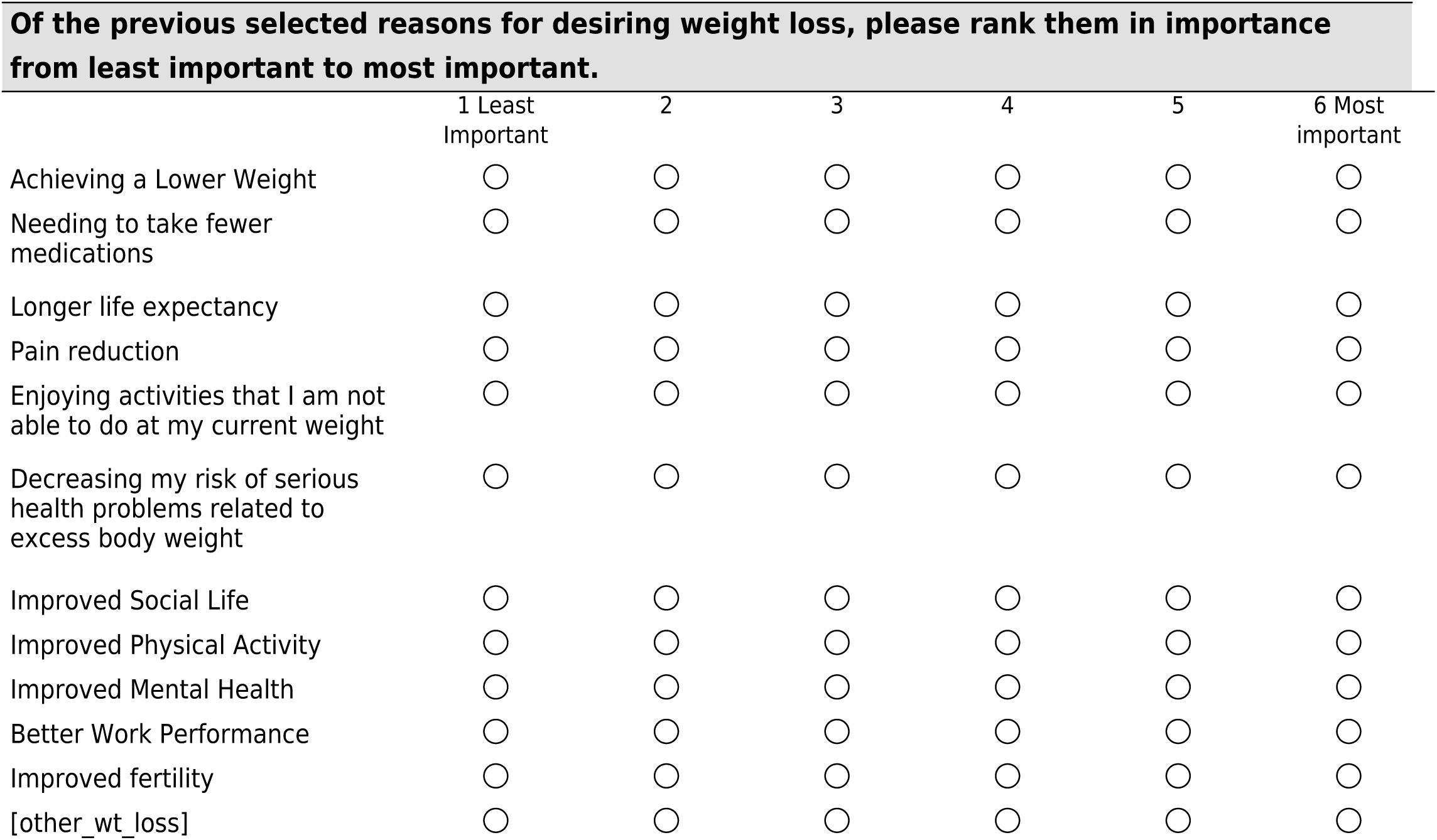
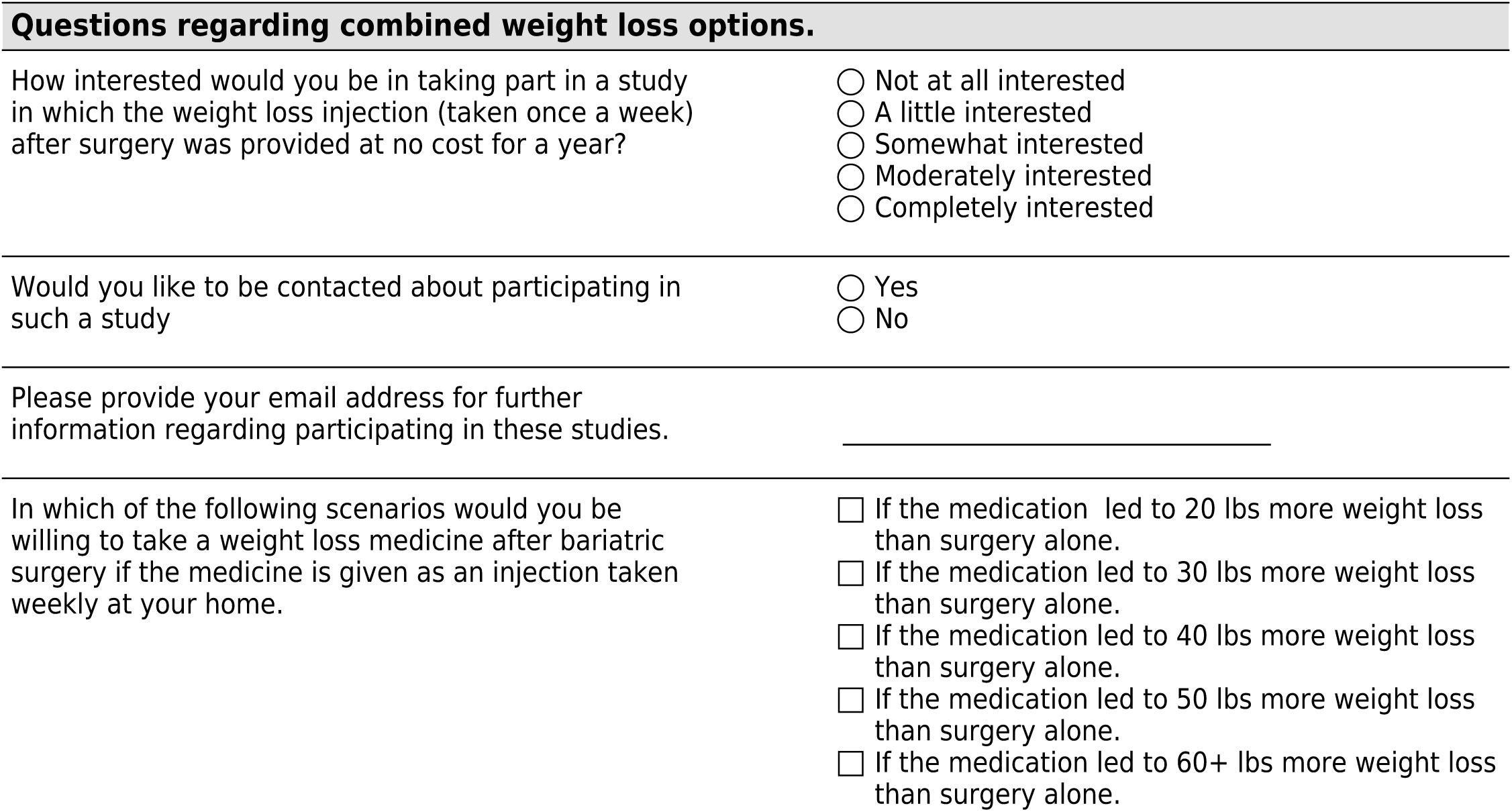
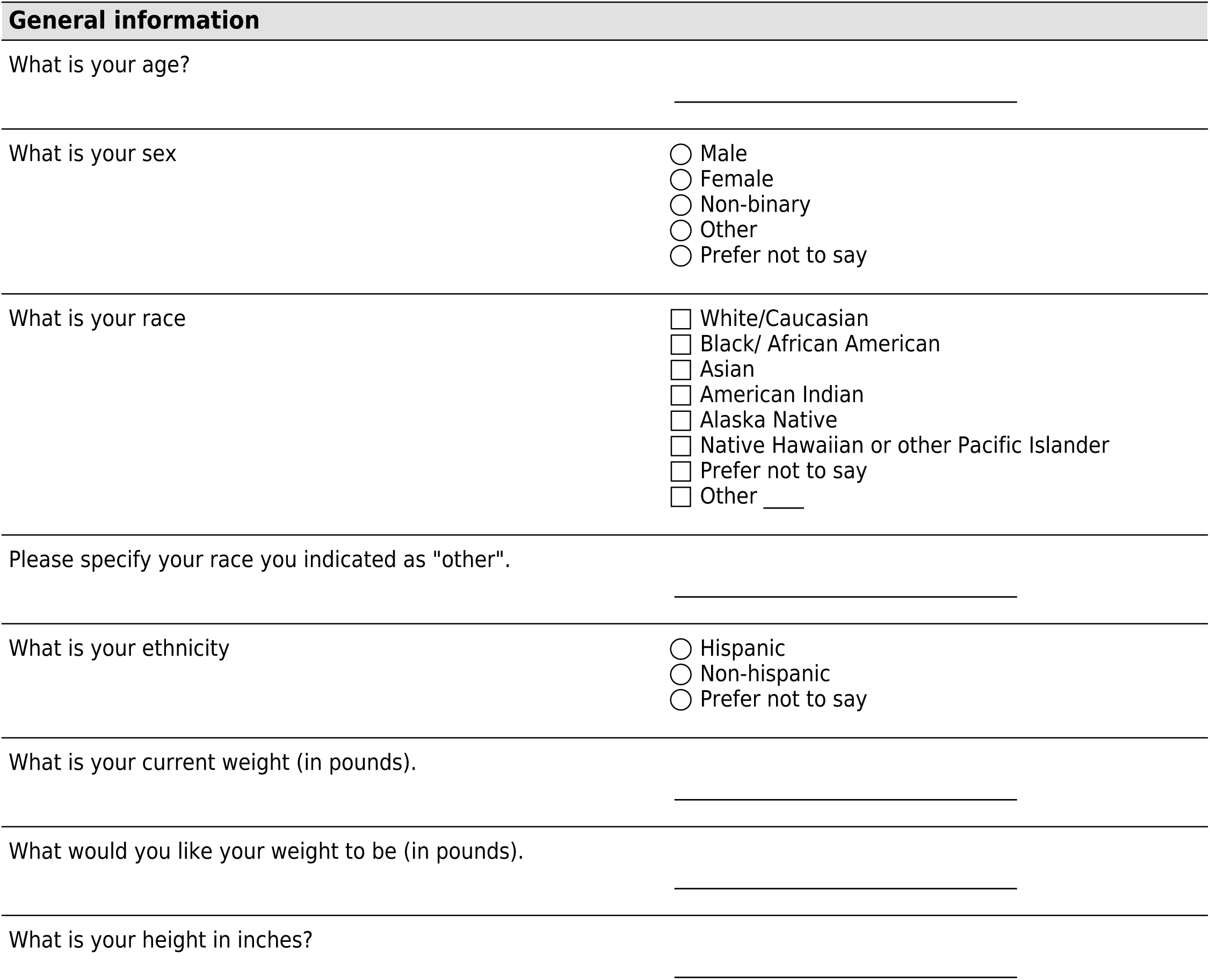
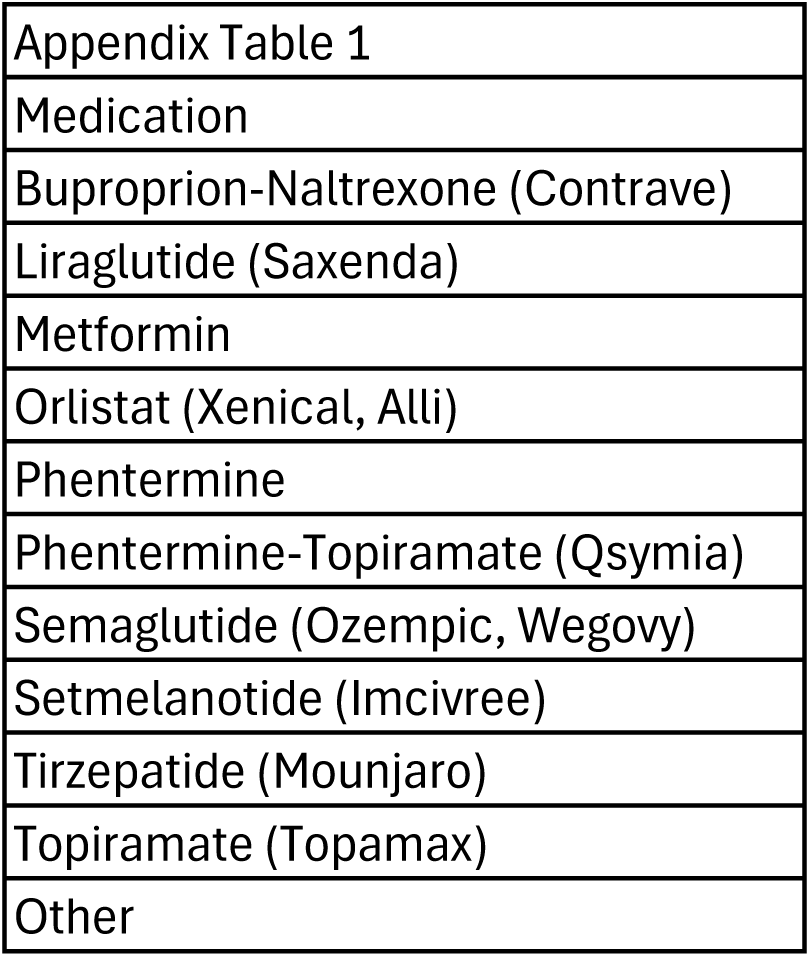
Comparison between patients with and without history of anti-obesity medication usage of selected motivations for seeking bariatric surgery.

We asked participants “How much weight (in pounds) do you (or did you) expect to lose with surgery?” The median response was 65.8 kg (IQR 56.7 – 77.7). This was on average 39% (IQR 34 – 47%) expected total body weight loss from bariatric surgery. Expected weight loss from bariatric surgery was similar among participants with or without a history of AOM use (68 kg vs 45 kg; 40% TWL vs 37% TWL, p=0.2).

## Discussion

We report a significant majority of patients have previously utilized AOMs prior to seeking surgical weight loss, with most patients trialing more than one AOM. The primary reason for seeking weight loss was most often to increase longevity and quality of life and was not associated with prior AOM use. More often participants reported a preference for surgical weight loss compared to medical but were willing to use AOMs to enhance weight loss after bariatric surgery.

The impact of preoperative weight loss, including through mandated medical weight loss programs, have been extensively investigated with minimal demonstrated benefit.(16) Interestingly, incorporation of AOMs as part of these preoperative weight loss programs are rare. This likely owes to the fact that while preoperative weight loss is encouraged, and occasionally even required prior to surgery, payers rarely cover AOMs as preparation for surgery.(16) The minimal existing literature on preoperative AOM use primarily details case reports of attempts at preoperative weight loss in patients in which at their current weight, bariatric surgery carries prohibitive risk.(17, 18) Our findings suggest that despite this absence of evidence guiding AOM use prior to surgical weight loss, use of AOMs prior to bariatric surgery appears to occur in a majority of patients seeking surgical weight loss. The majority of survey respondents reported use of AOMs prior to surgery and on average trialed two medications.

Several prior studies examined motivations and goals for weight loss as well, and our findings match closely with the current literature with some notable differences.(10, 14) In congruence with our study, prior publications have found improved overall health as the most common reason for seeking bariatric surgery.(11, 19, 20) However, in contrast to our findings, Ahlich found only 10% of patients reported improved longevity as a motivating factor for seeking surgical weight loss.(19) Hult et al.’s findings more closely matched our own, with weight loss, improved comorbidities, and longevity as the three most common motivators for weight loss, respectively in a multi-national, all female study.(12) To our knowledge, this is the first study to compare motivations for seeking bariatric surgery between patients who have and have not previously utilized AOMs. With the exception of patients’ desires to improve their physical activity, the two groups did not differ in stating the importance of the most common reasons listed for seeking surgical weight loss.

Our survey further reinforced the concept of expectations for surgery-induced weight loss exceeding the average weight loss achieved with sleeve gastrectomy and gastric bypass. This aligns with the current literature with expected excess weight loss of 72%-106%.(21–24) In our results, we report that patients were willing to utilize injectable AOM after surgery to achieve additional desired weight loss. Ultimately, more studies are needed to investigate the clinical and psychosocial benefits of enhanced weight loss with the use adjuvant medical therapies.

This study has several important limitations to consider. First, the sample of participants are those who have failed to achieve weight loss with AOMs and are interested and seeking bariatric surgery. Given that we did not have access to a non-surgical comparator group, the sample likely over represents those who failed medical treatments. Second, this study was conducted at a single surgical weight loss clinic with the respondent population predominantly white and female which may limit the generalizability. Third, the response rate for the survey was 32% of those contacted. Non-response bias may have influenced the results we report. We do not believe the sample of respondents were differential in their reported AOM usage prior to bariatric surgery, weight loss experiences, and how these experiences may have impacted motivations for seeking bariatric surgery. We did not have respondents expand on the duration of AOM use or ask about respondents compliance with AOMs. However, the average degree of weight loss reported suggests respondents utilized AOMs consistently and for a significant length of time. Lastly, we recognize that larger studies enrolling a more diverse sample from both medical and surgical clinics would add to and confirm these findings.

## Conclusion

AOMs are utilized by a majority of patients prior to seeking surgical weight loss, with most patients utilizing more than one AOM before considering surgery. With minimal data guiding AOMs in patients who are candidates for bariatric surgery currently, future studies should explore how use of these medications can be optimized to improve weight loss outcomes and enhance the effectiveness of surgery. Patients also continue to desire greater weight loss than is offered by surgery on average in isolation, and patients in our study expressed significant interest in utilizing AOMs after surgery to further enhance weight loss. Prospective studies and clinical trials are needed to determine the most effective way AOMs can be implemented after bariatric surgery.

## Data Availability

All data produced in the present study are available upon reasonable request to the authors

